# Environmental risk factors for self-harm during imprisonment: a prospective cohort study

**DOI:** 10.1101/2024.01.31.24302059

**Authors:** T Stephenson, I Harris, C Armstrong, S Fazel, R Short, N Blackwood

## Abstract

**Introduction:** Self-harm is a major public health issue in the imprisoned population. Limited high-quality evidence exists for the potential impact of prison environmental factors such as solitary confinement. This pilot prospective cohort study in a large male remand prison in England sought to estimate effect sizes of a comprehensive range of prison environmental factors on self-harming behaviours.

**Methods:** A random sample of all prisoners (*N*=149) starting a period of imprisonment at the study prison took part in a clinical research interview, which assessed a range of known risk factors for self-harm in prison. Information concerning environmental factors, including staff numbers, cell placement and movements, and engagement in work and activities were collected from prison records. Incidents of self-harm behaviour in the 3 months after entering prison were measured using medical records and self-report at end of follow-up. Multivariable logistic regression models were calculated individually for each predictor.

**Results:** Single cell placement (OR 4.31, 95% CI 1.06-18.24, p=0.041) and more frequent cellmate (OR 1.52, CI 1.14-2.17, p=0.009) and cell (OR 1.83, 95% CI 1.28-2.86, p=0.003) changes were associated with an increased risk of self-harming behaviour. Unexpectedly, a lower staff-to-prisoner ratio (OR 0.89, CI 0.78-0.99, p=0.039) was also associated with an increased risk of self-harming behaviour. Following sensitivity analyses, the associations between frequent cell changes and self-harm behaviour, and between single cell placement and self-harm ideation, remained statistically significant.

**Discussion:** This pilot study provides prospective longitudinal data regarding relationships between prison environmental factors and self-harm behaviour. Findings regarding single cell accommodation and frequent cell changes are consistent with the prior evidence base largely derived from case-control study data. The finding regarding frequent cellmate changes predicting self-harm is novel. Findings regarding staff-prisoner ratio and self-harm most likely reflect a reverse causal relationship. Replication in larger cohort studies is required to address the limitations of this pilot study.

## Introduction

Self-harm is a major public health issue in prisons. Rates of self-harm in England and Wales have more than doubled in the last ten years, from less than 300 per 1,000 prisoners in 2013 to 805 per 1,000 prisoners in the 12 months to September 2023^1^. Self-harm behaviour in prison is an established risk factor for subsequent suicide, both in prison and on release into the community^2,3^. The assessment and management of self-harm in prisons is a significant resource burden for prison and healthcare services.

A recent systematic review and meta-analysis highlighted the empirical evidence for a range of individual characteristics contributing to self-harm behaviour in prison populations, including sociodemographic, criminological, historical, clinical and psychosocial variables^4^. This study identified moderate-sized effects on self-harm for prison environmental factors such as exposure to solitary confinement, single cell placement, lack of social contact, and lack of employment in prison. One case-control study additionally identified a small effect on self-harm behaviour from frequent cell moves within prison over the previous two years (OR 1.08)^5^.

However, there is limited high-quality evidence regarding the prospective longitudinal relationship between the kind of events, experiences and environments encountered by prisoners and their self-harming risk. A prognostic model developed to help risk assessment in the area does not include any prison environmental variables^6^. Recent work reviewing the available evidence has suggested that prison regime characteristics, namely time out of cell and time in purposeful activity, may influence self-harm and suicide risk in prisons^7^. In addition, qualitative and cross-sectional research implicates a broader range of prison environmental factors potentially implicated in self-harm such as staffing levels, relationships between prisoners and staff, staff turnover, prisoner turnover and overcrowding^8-12^. However, the strength of such potential environmental effects is currently unclear. Longitudinal study designs could provide high-quality evidence regarding these relationships, which could in turn inform the clinical assessment and management of self-harm in this population and suicide prevention strategies in prisons.

The aim of this pilot prospective cohort study was therefore to estimate the effect sizes of potentially relevant prison environmental variables on self-harm behaviour in the male prison population. A secondary aim was to inform a power calculation for a larger longitudinal study of the effects of individual and environmental exposures on self-harm behaviour in the same population. The study was part of a wider mixed methods project which included qualitative work informing the choice of exposure variables.

## Methods

### Sample

We carried out a 3-month prospective cohort study of new entrants to a large Category B men’s prison in London, England. Category B prisons in general have a greater emphasis on security and less emphasis on training and rehabilitation than those in Category C and D. The study prison is a “local” prison housing prisoners taken directly from courts in the local area. The population of the prison is largely on remand (awaiting trial) but has sizeable minorities of sentenced prisoners and people detained under immigration law. Previous longitudinal research at the same study site documented a higher-than-average prevalence of self-harm behaviour making it well suited for our study. We chose a short follow-up period of three months due to the high attrition rates inevitably found in a remand prison^13^.

The study population consisted of all male prisoners who had recently arrived (within one week of sampling) into the study prison between March and July 2022. Potentially eligible subjects were identified from prison reception lists via the National Offender Management Information System (NOMIS). A simple random sample of subjects was taken each week, totalling 675 across the duration of the study (see Figure 1). 420 of those sampled met initial eligibility criteria (aged 18 or older and current prison spell lasting less than one month). We approached potential participants on prison wings to carry out screening based on further eligibility criteria and to obtain informed consent. Subjects were excluded if their English language skills were insufficient or if they lacked decision-making capacity to give informed consent. 152 participants were recruited to the study. Following a baseline assessment interview, three participants withdrew their consent leaving a baseline cohort of 149 participants. Participants were followed up for a mean duration of 73.5 days, with 83 participants (55.7%) completing the 3-month follow-up period. 66 participants (44.3%) were lost-to-follow-up due to release or transfer to another prison. Four participants completed follow-up but did not complete the exit interview. As this was an exploratory pilot study, the sample size was determined by available resources rather than by a power calculation.

**Figure 1.**
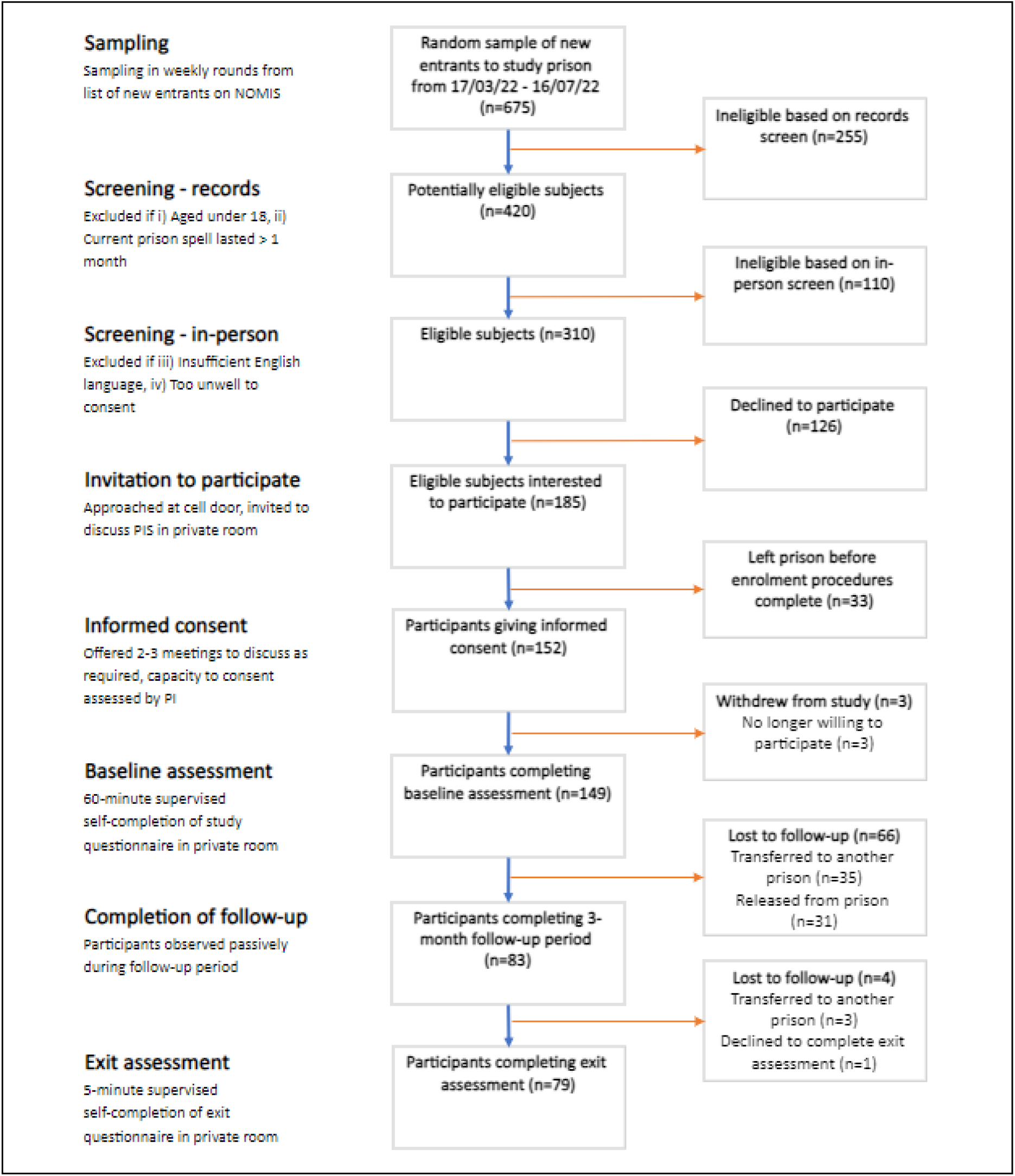
Study flow chart.

Ethical approval included permission to access non-identifiable demographic data (ethnicity and age) of non-participants, allowing for comparison of those who participated (n=149) and those who were eligible but declined participation (n=126, 41%). There were no identifiable differences between these groups in terms of ethnicity (χ^2^ = 2.86, df = 4, p = 0.582) or calendar age (*t* = 0.097, p = 0.923).

### Procedure and measures

Participants took part in a total of 1-3 meetings with researchers, as required, before giving written consent. Verbal consent was documented for those without sufficient literacy skills (n= 1). All screening procedures were carried out by TS, a psychiatrist, who assessed capacity to consent. Subsequent consent and baseline interview procedures were carried out either by TS or by IH.

Recruitment and interview procedures were carried out within approximately six weeks of arrival into custody (range 4-42 days, mean 25.6 days). A baseline interview assessed the presence of a range of self-harm risk factors identified from the literature. Validated measures included the Drug Use Disorder Identification Test (DUDIT), the Childhood Trauma Questionnaire (CTQ), and the Barrett Impulsiveness Scale (BIS-II). Where validated measures were not available, questionnaire items were adapted from a previous study questionnaire (OxSHIP) assessing self-harm risk factors on entry to prison^6^. Other items were added to this to form an adapted 30-item SHAPE study questionnaire (see supplementary materials). Other demographic data, including ethnicity category, were collected from the NOMIS record. Due to expected low literacy levels in the study population, all questionnaires were administered in interview format by TS and IH.

Putative prison-specific risk factors were identified by scoping the existing literature and from preliminary findings of a parallel qualitative study by the authors of the perspectives of prisoners and staff on events and experiences in prison affecting self-harm behaviour (in preparation). Operational definitions for 15 exposure factors were refined iteratively by the study team alongside scoping of exposure data availability and format at the study site. These included measures of prisoner activity (time in activities and work status), disciplinary measures (incentive level), cell-related factors (solitary confinement placement, single cell placement, number of cellmates, number of cell changes), contact with the outside world (social visit status, days to first phone call, time on phone), measures of staff and peer support worker numbers (staff-prisoner ratio, peer listener-prisoner ratio) and of staff responsivity (emergency bell response rate) and miscellaneous measures (vape use and COVID-19 isolation measures).

Three exposure variables (emergency bell response rate, staff-prisoner ratio and peer listener-prisoner ratio) were assessed by linking local group-level data for each prison wing with participants’ starting location for each study week. Running weekly mean values were calculated from daily data for each variable. Measures of phone use were assessed using a British Telecom prison phone database. We identified COVID-19 isolation from a local study prison dataset. Vape use was assessed by self-report at baseline interview. All other exposures were measured using routinely collected data on participants’ NOMIS prison record. Table 1 details operational definitions and data characteristics for all study variables.

**Table 1.** Operational definitions and data characteristics for exposure, confounder and outcome variables.

| Exposure variable | Definition | Units | Data type | Data source | Completeness in % (*count*) |
| --- | --- | --- | --- | --- | --- |
| Time in activities | Total time in any prison activity, including courses and education but excluding work | Days | Continuous | NOMIS | 100 (82/82) |
| Working status | Any days in an allocated job in prison | - | Binary | NOMIS | 100 (82/82) |
| Incentive level <sup>1</sup> * | Prisoner incentive level during follow-up (Standard-throughout, Ever-enhanced, Ever-basic) | Category | Categorical | NOMIS | 92.7 (76/82) |
| Solitary confinement | Any placement in prison's Care & Separation Unit (solitary confinement) during follow-up | - | Binary | NOMIS | 100 (82/82) |
| Single cell <sup>2</sup> | Any placement in a single occupancy cell | - | Binary | NOMIS | 95.1 (78/82) |
| Cell changes * | Total number of cell changes | - | Continuous | NOMIS | 100 (82/82) |
| Cellmates * | Total number of cellmates | - | Continuous | NOMIS | 94.9 (74/78) |
| Social visit | Receipt of any social visit | - | Binary | NOMIS | 86.6 (71/82) |
| Time to first phone call <sup>3</sup> * | Time from prison entry to first phone call using permanent PIN | Days | Continuous | Prison phone database | 96.3 (79/82) |
| Time on phone * | Total time spent on phone using permanent PIN (excluding phone calls lasting less than 30 seconds) | Hours | Continuous | Prison phone database | 57.3 (47/82) |
| Vape use * | Any current vape use at time of baseline assessment (self-report) | - | Binary | Self-report (baseline) | 98.7 (81/82) |
| COVID-19 isolation <sup>6</sup> * | Any individual COVID-19 isolation (whether originating from participant or cellmate infection) | - | Binary | Local prison data | 100 (82/82) |
| Emergency bell response rate <sup>4</sup> * | Daily proportion of emergency bell calls that are answered within target time of 5 minutes (assessed at wing-level, linked to participants' location) | % | Continuous | Local prison data | 100 (214/214) |
| Staffing ratio | Daily mean staffing number in morning / evening shifts, per wing capacity (assessed at wing-level, linked to participants' location) | Ratio | Continuous | Local prison data | 99.5 (213/214) |
| Prison Listener ratio <sup>5</sup> | Daily mean Prison Listener number, per wing capacity (assessed at wing-level, linked to participants' location) | Ratio | Continuous | Local Samaritans data | 100 (214/214) |
| **Confounder variable** |  |  |  |  |  |
| Age |  | Years | Continuous | NOMIS | 100 (82/82) |
| Ethnicity | National Offender Management Information System (NOMIS) ethnicity category: White, Black, Asian, Mixed, Other | Category | Categorical | NOMIS | 100 (82/82) |
| Violence history | Defined as any previous conviction for a violent offence | - | Binary | Self-report | 100 (82/82) |
| **Outcome variable** |  |  |  |  |  |
| Self-harm episode | Any episode of self-harm behaviour | - | Binary | Prison medical record (Systm1) | 100 (82/82) |
|  |  |  |  | Self-report (exit interview) | 95.1 (78/82) |
| Self-harm ideation | Any thoughts of self-harm behaviour | - | Binary | Self-report (exit interview) | 95.1 (78/82) |

<sup>1</sup> The Incentive and Earned Privilege Scheme (IEPS) is a behavioural incentive programme used across prisons in England and Wales. New prisoners start at 'standard' level. Downgrading to 'basic' for results in loss of earnable incentives including extra and improved visits; eligibility to earn higher rates of pay; access to in-cell television; opportunity to wear own clothes; access to private cash and additional time out of cell. It is separate from the disciplinary system.
<sup>2</sup> Double cell occupancy is standard in the study prison. Single cell occupancy in 'ordinary location' (i.e. outside of solitary confinement or healthcare wing) can result from different scenarios including high violence risk rating by the prison, medical reasons (for example, severe incontinence or severe PTSD) or temporarily whilst awaiting a new cellmate.
<sup>3</sup> New prisoners are allocated a Personal Identification Number which is used to make out-going phone calls from the cell. A contact must be approved by the prison prior to the first phone call.
<sup>4</sup> Emergency cell bells are available in each prison cell for prisoners to seek urgent attention from an officer. The target response time in the study prison is 5 minutes.
<sup>5</sup> The Listener scheme is a peer-support scheme within prisons run by Samaritans, a large suicide prevention charity. Listeners are trained by volunteers to provide confidential emotional support for peers who are struggling to cope or feeling suicidal.
<sup>6</sup> In the study prison, isolation protocols in place during the study period stated that in cases of any confirmed COVID-19 infection amongst occupants of a double occupancy cell, both occupants should isolate for 10 days.
\* These variables were included based on preliminary findings from parallel qualitative work.

The primary study outcome was any self-harm episode during the 3-month follow-up, assessed from both participants’ prison medical record and by semi-structured interview at end of follow-up. The outcome was double-assessed to test reliability of records-based assessment for future research. Self-harm was defined as any intentional self-poisoning or injury irrespective of the apparent purpose of the act^14^. The secondary outcome was any self-harm ideation during follow-up, assessed by semi-structured interview at the end of follow-up. This was included with a view to improving statistical power as it was expected to be more prevalent than self-harm episodes.

The exit interview was carried out by TS within approximately 4 weeks of completion of 3-month follow-up (range - 4-31 days, mean 7.7 days). Wording of the exit interview items was adapted from items in the Self-Injurious Thoughts and Behaviours Interview (SITBI)^15^, accounting for a broader definition of self-harm as above (see supplementary materials).

Ethical and HMPPS regulatory approval for the study were given by NHS (22/WA/0007) and HM Prison and Probation Service (2022-009).

### Statistical analysis

Those lost-to-follow-up (n=66) were excluded from analyses. We excluded participants with any placement in the healthcare wing (n=1) from all analyses because of its markedly different environment which resulted in outlier values for many exposure variables, for example, staffing ratio. The final prospective cohort thus consisted of 82 participants.

We categorised one exposure variable – incentive level – where all participants fell into one of three categories (ever basic, standard throughout, ever enhanced). Participants with missing data for individual-level exposures were excluded from each analysis (see data completeness, Table 1). One missing group-level exposure data point for staff-prisoner ratio was imputed by calculating the mean of values for the previous and forthcoming weekdays. Reliable occupancy data were not available to calculate staff-prisoner ratio and peer listener-prisoner ratio. Therefore, total capacity was used as a proxy assuming 100% occupancy throughout the study. One prison wing was closed at the start of the study but gradually reopened during the second half of the study and reached approximately 50% capacity at end of the follow-up period. The total occupancy for this particular wing was imputed as 25% of capacity throughout the period of reopening based on consultation with prison officials.

We described the data using frequencies and proportions for categorical variables, means and standard deviations for normally distributed numerical variables, and medians and inter-quartile ranges (IQR) for discrete and skewed numerical variables.

Odds ratios were calculated for the association of each prison-specific exposure with both study outcomes using separate multivariable logistic regression models in R. Due to low- or zero-counts for both study outcomes, we did not carry out analyses for solitary confinement as a predictor.

Adjustments were made for participant calendar age, ethnicity (white/non-white), and violent offending history. These factors were pre-selected as confounders based on their theoretical impact on both exposure and outcome. In a logistic regression model containing only presumed confounders and the study outcomes, white ethnicity was found to strongly predict self-harm episode (OR 11.41, CI 2.07-213.73, p=0.023) but not self-harm ideation (OR 1.97, CI 0.66-6.42, p=0.233), whilst an association was seen between violent offending history and self-harm ideation (OR 2.62, CI 0.93-7.75, p=0.071) but not self-harm episode (OR 2.05, CI 0.59-7.80, p=0.269). No significant associations were seen between age and either outcome. A p-value cut-off of <0.05 was used to designate statistical significance for all tests.

We carried out sensitivity analyses for outlier values, defined as >3 SD from the mean. Outliers existed for three continuous variables, number of cell changes (n=2), number of cellmates (n=1) and staff-prisoner ratio (n=1). A separate sensitivity analysis was carried out for all predictors by removing participants with any placement in solitary confinement (n=2) from all analyses.

## Results

### Cohort characteristics

Cohort demographics are described in Table 2. Participants (n=149) were all men with a median age of 34 (IQR 27-43). The sample was representative in age (χ^2^ = 2.82, df = 3, p=0.421) and ethnicity (χ^2^ = 3.74, df = 4, p=0.443) of the wider population at the study prison based on data from 2021^16-17^. Those who completed follow-up were comparable to those lost-to-follow-up except that they were more likely to have been on remand at time of baseline assessment. The majority of participants were on remand at time of entry to the study (74.5%). A large proportion of prisoners who were already sentenced at entry to the study (21/25, 84%) were lost to follow-up due largely to transfer to other prisons. A previous conviction for violent offence (68/149, 46.3%) and an active diagnosis or treatment for any mental disorder at baseline interview (59/149, 39.9%) were self-reported large minorities of the cohort, respectively. 40/149 (26.9%) of participants reported a history of self-harm outside of prison.

**Table 2.**
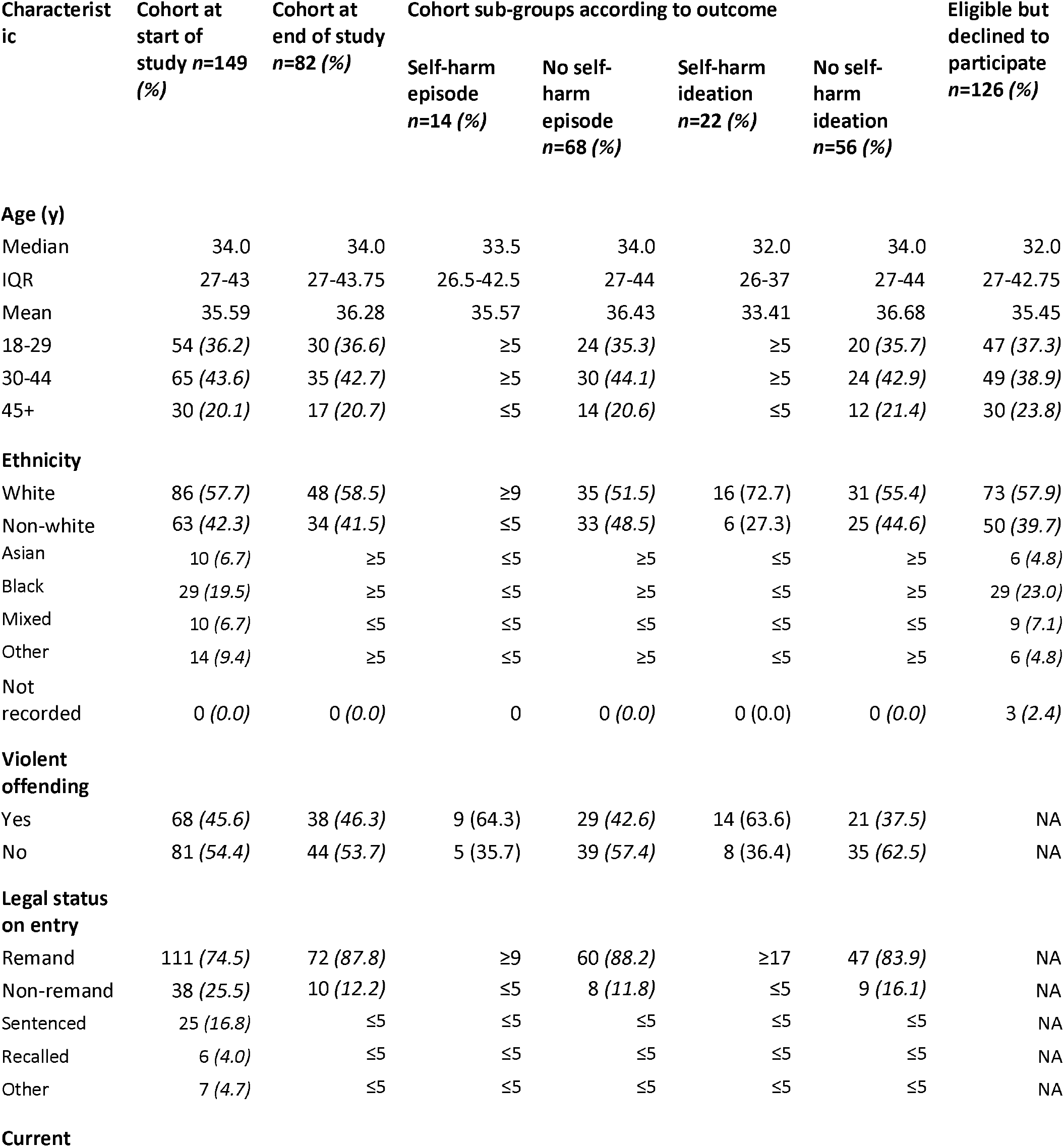
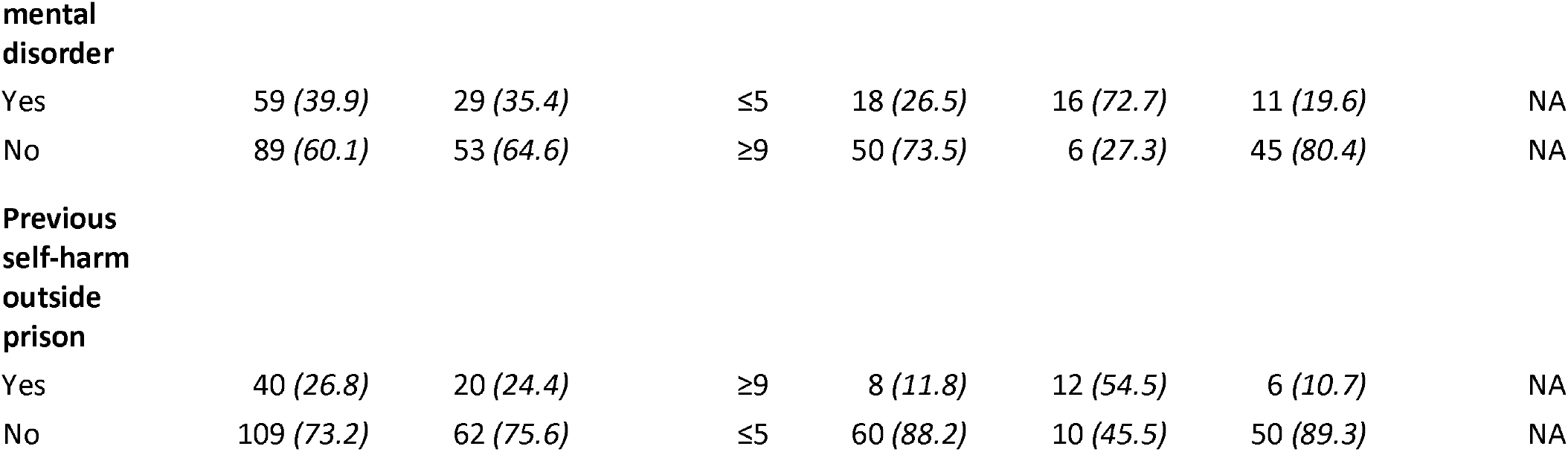
Cohort characteristics at beginning and end of study compared with those declining to participate.

The event rate for any self-harm episode from records during follow-up was 17.1% (14/82). This was comparable to the rate assessed from self-report of 16.7% (13/78; see supplementary materials). The corresponding rate for any self-reported self-harm ideation was 28.2% (22/78).

Outcome counts for categorical and binary exposure variables are shown in Table 3. Table 4 presents median and IQR values for skewed numerical exposures and mean and SD values for normally distributed exposures. There were missing values for incentive level (n=6), cellmates (n=4), single cell placement (n=4), social visit (n=11), first phone call (n=3), total phone use (n=35) and vape use (n=1).

**Table 3.**
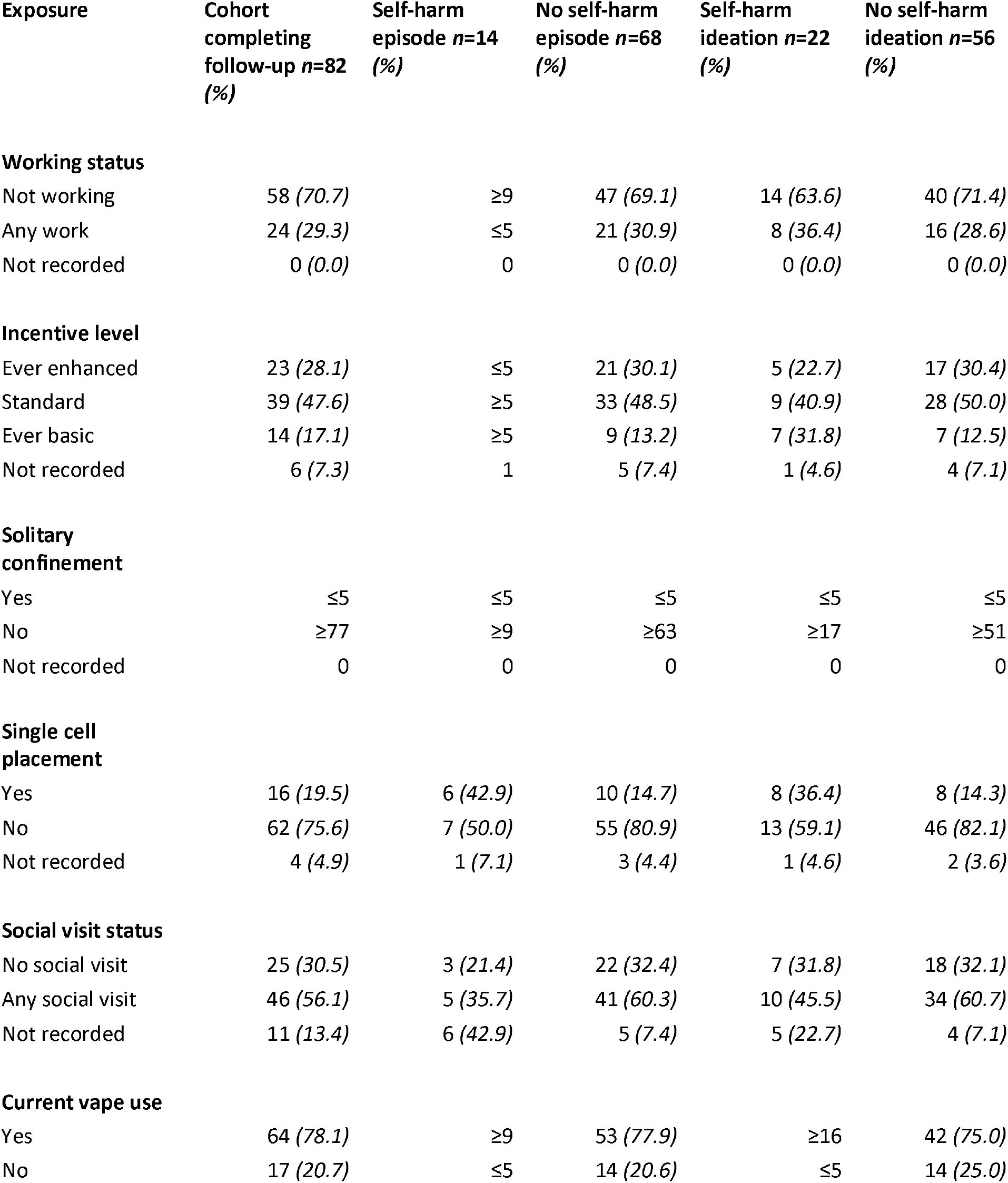
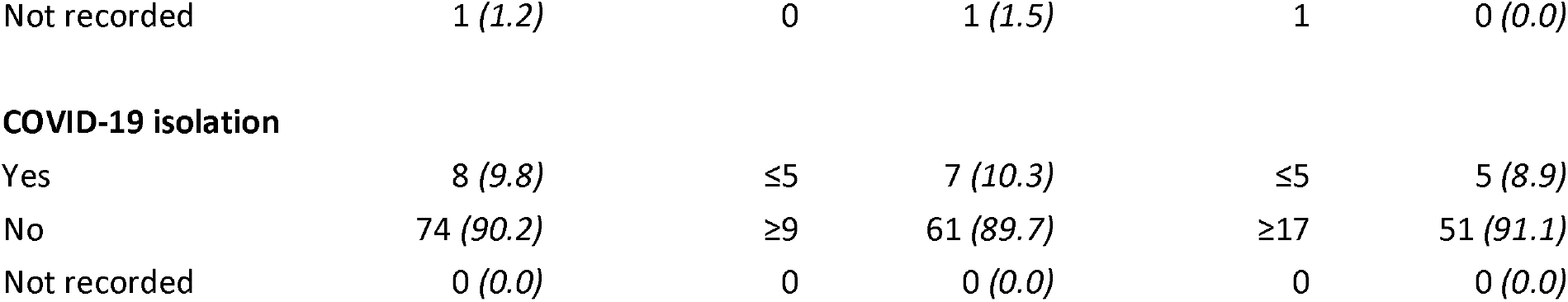
Categorical and binary exposures and self-harm outcome frequencies. Data not presented on distribution of exposure values between self-harm ideation outcome groups for participants where this outcome is missing (n=4).

**Table 4.**
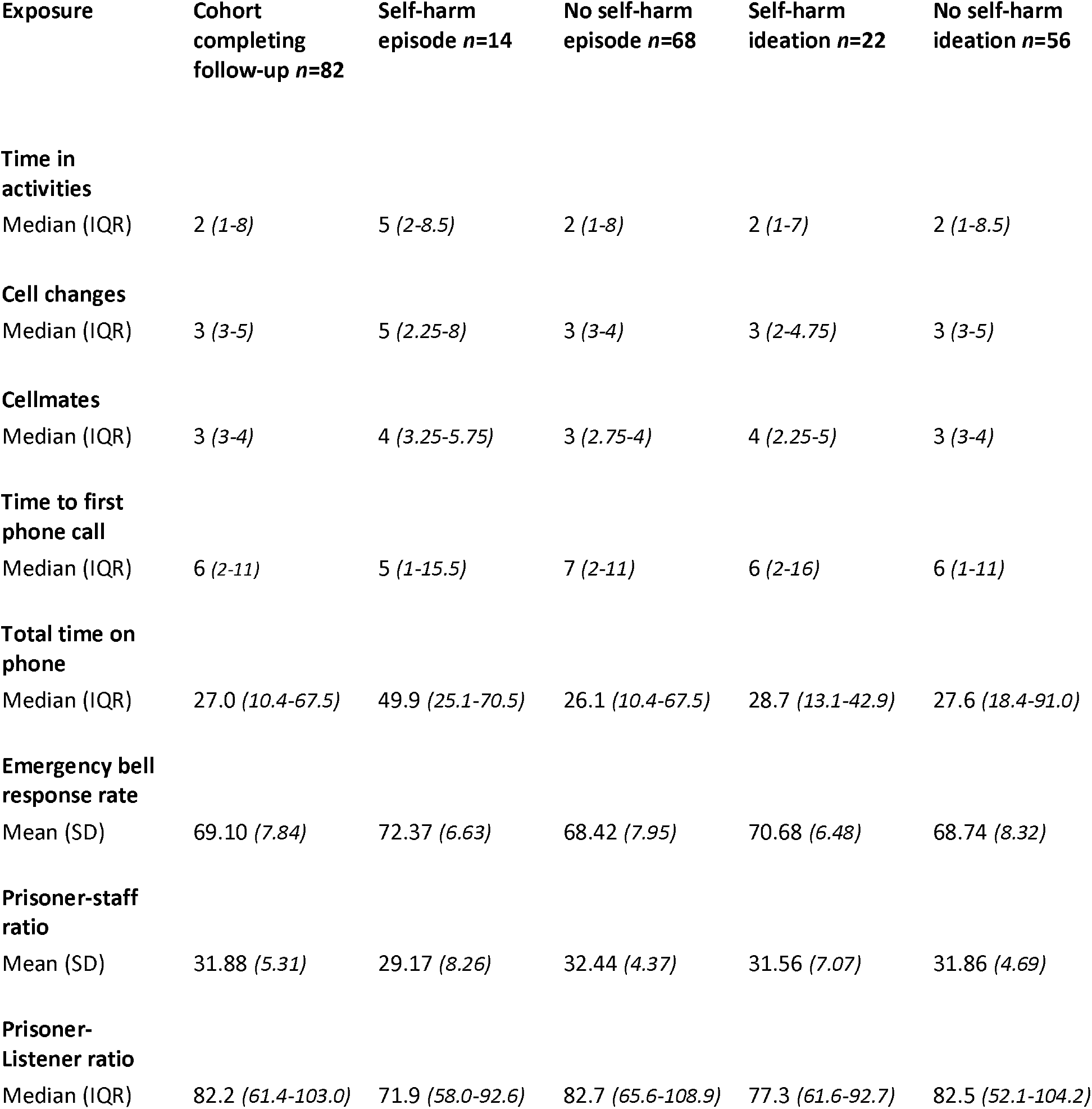
Continuous exposure distributions according to outcome. Data not presented on distribution of exposure values between self-harm ideation outcome groups for participants where this outcome is missing (n=4).

### Regression analyses

Adjusted and unadjusted odds ratios for logistic regression models are shown in table 5. More frequent cell changes (OR 1.83, CI 1.28-2.86, p=0.003), a higher number of different cellmates (OR 1.52, CI 1.14-2.17, p=0.009), and any placement in a single cell (OR 4.31, CI 1.06-18.24, p=0.041) were significantly associated with self-harm behaviour in adjusted models. A higher number of prisoners per member of staff was significantly associated with reduced self-harm behaviour in adjusted models (OR 0.89, CI 0.78-0.99, 0.039). There was no significant relationship between self-harm episode and other variables such as time in activities, non-working status, a lack of social visits, time to first phone call, total phone time, vape use or COVID-19 isolation.

**Table 5.**
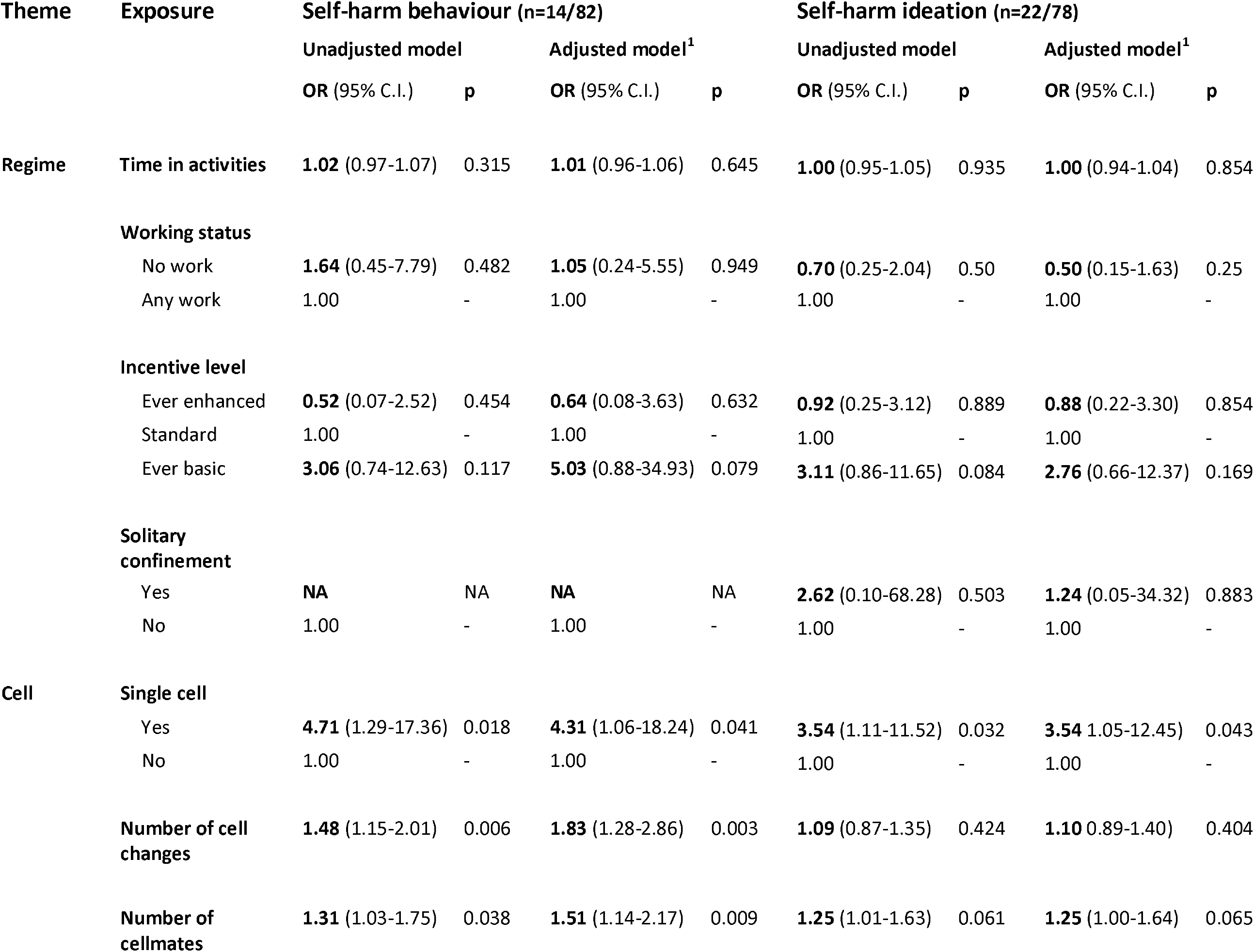
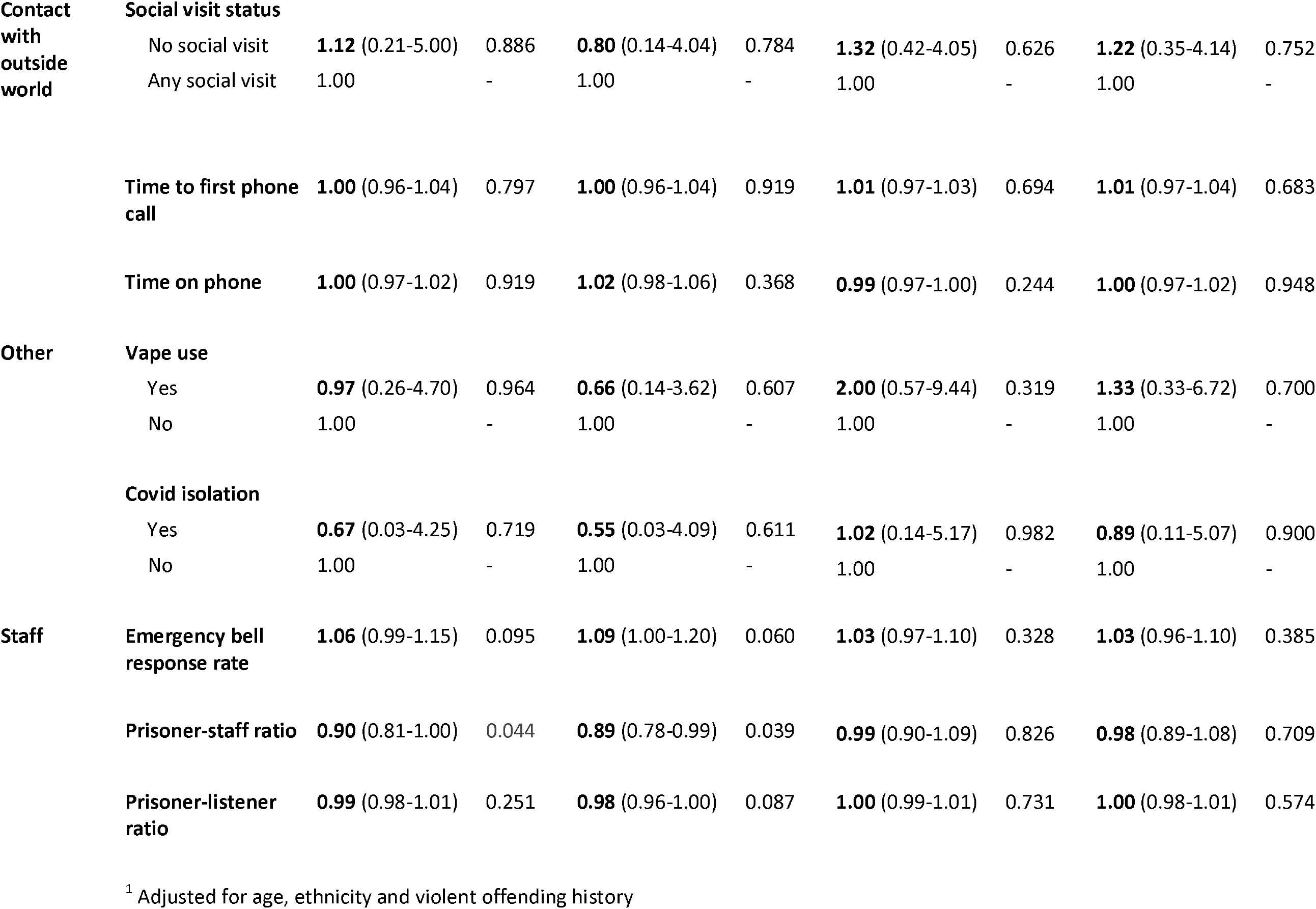
Odds ratios of predictors for self-harm episode and ideation in adjusted and unadjusted logistic regression models.

Single cell placement (OR 3.54, CI 1.05-12.45, p=0.043) was associated with self-harm ideation in adjusted logistic regression models (see table 5). No relationship was identified with the remaining variables and self-harm ideation.

### Sensitivity analyses

The significant association between more frequent cell changes and self-harm episode remained after excluding two high outlier values. However, the association between more frequent cellmate changes and self-harm episode was no longer significant after excluding one high outlier value (OR 1.33, CI 0.96-1.96, p=0.112). The association seen for lower staff-prisoner ratio and self-harm episode was no longer significant after excluding one low outlier value (OR 0.97, CI 0.83-1.15, p=0.734).

After excluding participants with any placement in solitary confinement, the association seen between self-harm episode and single cell placement (OR 3.01, CI 0.63-13.7, p=0.150) was no longer significant. No other associations seen with self-harm episode were significantly affected by excluding such participants. The association between single cell placement and self-harm ideation remained significant after excluding participants with any placement in solitary confinement.

## Discussion

In this study of 149 prisoners, we tested 15 measures of environmental risk factors with self-harm over a 3-month period in one male English prison. We found that more frequent changes of cell and cellmate and single cell placement were associated with a higher risk of self-harm behaviour. Unexpectedly, a higher number of prisoners per staff member was associated with a lower risk of self-harm behaviour.

A recent meta-analysis of case-control and cross-sectional evidence suggested that custody-specific factors may confer increased risk of self-harm^4^. These included aspects of prisoners’ relationships with staff and peers (disciplinary infractions, sexual or physical victimisation, being threatened with violence), cell placement (solitary confinement), and a lack of work and activities and social contact or visits in prison. However, we are not aware of longitudinal studies that have examined such relationships, except in the cases of solitary confinement^18-20^ and disciplinary infractions^20-21^. This pilot prospective cohort study is the first longitudinal study, to our knowledge, that examines the effects of various environment factors on self-harm behaviour in prisons including single cell placement, cell changes, and measures of work and activities. It is also novel in investigating the effects on self-harm of incentive level, number of cellmates, measures of in-cell phone use, and measures of prison staffing level, staff responsiveness to prisoners and proximity of peer listeners.

The association seen between self-harm and frequent cell changes accords with findings from one case-control study in which high housing (prison cell) moves in the previous two years was associated with self-harm^5^. Our finding regarding single cell placement is in keeping with the findings of a recent meta-analysis^4^ which identified a similar sized effect on self-harm behaviour (OR 1.5, 95% CI 0.8-2.9) but which was based on case-control and cross-sectional data. The association between self-harm episode and greater number of cellmates is novel, and similar in size to that seen for cell changes. The finding that lower staff-prisoner ratio was associated with increased odds of self-harm episode is novel and unexpected.

The lack of significant associations between self-harm outcomes and time in activities, working status and social visit status may be due to the sample being underpowered. Previous studies have identified a link between these variables and self-harm, including the results from a large meta-analysis regarding not working in prison (OR 1.9, 95% CI 1.5-2.5, n=3311) and no social contact or visits (OR 2.3, 95% CI 1.5-3.5, n=2153)^4^, and an ecological study of UK prisons which identified an association between prisons with lower time in purposeful activity and completed suicide amongst prisoners^22^.

### Limitations

The small sample size of this study meant that our analysis was underpowered to detect small-moderate effects. It also reduces the precision of effect size calculations. Results for cell changes, number of cellmates and staff-prisoner ratio were sensitive to the impact of outlier exposure values, and the result for single cell placement was sensitive to removal of the sub-group with solitary confinement placement.

Aggression is a plausible confounder of the relationship between prison environment factors and self-harm, with prisoner aggression determining single cell placement, number of cell changes and cellmates as a result of prison cell sharing risk assessments (which are concerned with the risk of inter-prisoner aggression, see Table 2 footnotes). We controlled for previous violent offending but did not measure in-study aggression amongst participants. Unmeasured confounding from in-study aggression may explain the loss of significance of the relationship between single cell placement and self-harm episode when excluding the sub-group who also had solitary confinement placement. The direction of any bias introduced by unmeasured confounding is unclear, but its magnitude may have been reduced by adjusting for previous violent offending status given that this predicted self-harm ideation amongst the cohort.

The large proportion of participants lost-to-follow-up in this study (44.3%) raises the risk of selection bias in those completing follow-up. Reassuringly, completers were comparable in age and ethnicity category to the original cohort. Similarly, this study used records to measure exposures and outcomes, raising the risk of information bias. However, primary outcome assessment using prison medical records and self-report produced highly similar results.

The timing of exposure and outcome were not measured in this study, so it was not possible to demonstrate the direction of effect in the relationships identified between environmental factors and self-harm outcomes. Reverse causality may explain the association of self-harm episode with lower staff-prisoner ratio, given the common practice of monitoring prisoners who self-harm with regular or constant staff observation, resulting in higher staffing levels for the surrounding area.

A small sample size prevented us from assessing the extent and impact of collinearity between predictors. High collinearity between multiple environment predictors is plausible, for example between participants’ cell changes and total number of cellmates. We were also unable to assess whether any other known risk factors for self-harm, such as mental disorder, psychological distress and history of abuse mediated or moderated the effects of the study exposures. One plausible explanation of our cell-related findings is that they are mediated by unmeasured individual differences in problems with emotion dysregulation and/or aggression.

### Future research

The relationships identified would benefit from replication in better powered longitudinal studies. Basic incentive level and emergency bell response rate exposures warrant further examination in larger samples given that the results in this small sample approached the threshold for statistical significance. Larger samples will also permit sub-group analyses. Future studies using survival analyses approaches could discern the direction of relationships observed. Replication of the study findings in research in prisons of varying security, location and function, and in the female prison population, will improve generalisability. Future research should seek to address in-study aggression as a potential confounder of prison environment exposures and self-harming outcomes. The high similarity of outcome event rates assessed via prison medical records and self-report may have implications for future study designs in this area.

We identified 12 other potentially relevant environment factors which could not be assessed in this study due to non-availability or poor quality of data including time out of cell, prison regime cancellations, bullying and victimisation and prison occupancy rate. Where feasible, future research should attempt to examine the relationships of such factors with self-harm.

### Clinical implications

If replicated, the relationships observed in this study between self-harm and several prison environment factors may have implications for the assessment and management of self-harm by clinical and non-clinical staff. The association between single cell placement and self-harm highlights the importance of approaches prisons can take to reduce morbidity and mortality associated with self-harm, including – where appropriate – placement in safe (ligature-free) cells and the use of non-ligature clothing.

## Supporting information

S1 Baseline questionnaire

S2 Exit questionnaire

## Data Availability

Data cannot be shared publicly due to concerns that, even once anonymised, participants in this vulnerable population might be identifiable given low counts for many study variables.
Requests for study data can be directed to the HM Prison and Probation Service National Research Committee.

## Roles of researchers

TS was Chief Investigator for the project and was responsible with NB for study design. TS carried out all screening and initial approach procedures, counselled potentially eligible subjects about the study and assessed decision-making capacity. IH and CA were graduate level MSc psychology candidates who carried out baseline interviews and records data extraction alongside TS. RS and SF provided advice on the statistical analysis, which TS carried out. NB provided overall supervision for the project. All authors contributed to the manuscript.

## Notes

### Competing Interest Statement

The authors have declared no competing interest.

### Funding Statement

TS received funding for this research under a Preparatory Clinical Research Training Fellowship from the NIHR Maudsley BRC (IS-BRC-1215-20018). SF is funded by the NIHR Oxford Health BRC.
The funders had no role in study design, data collection and analysis, decision to publish, or preparation of the manuscript.

### Author Declarations

Ethical and HMPPS regulatory approval for the study were given by NHS Wales 3 Research Ethics Committee (22/WA/0007) and HM Prison and Probation Service National Research Committee (2022-009).

## References

1. Ministry of Justice. Safety in custody statistics, England and Wales: Deaths in prison custody to December 2023 assaults and self-harm to September 2023. 2024 [Available online at https://www.gov.uk/government/statistics/safety-in-custody-quarterly-update-to-september-2023/safety-in-custody-statistics-england-and-wales-deaths-in-prison-custody-to-december-2023-assaults-and-self-harm-to-september-2023, accessed 29/01/24.]

2. Hawton K, Linsell L, Adeniji T, Sariaslan A and Fazel S. Self-harm in prisons in England and Wales: an epidemiological study of prevalence, risk factors, clustering, and subsequent suicide, Lancet, 2014;383(9923) pp1147–54.

3. Zhong S, Senior M, Yu R, Perry A, Hawton K, Shaw J, Fazel S. Risk factors for suicide in prisons: a systematic review and meta-analysis, Lancet Public Health 2021;6(3):e164–e174. doi: 10.1016/S2468-2667(20)30233-4. Epub 2021 Feb 10. PMID: 33577780; PMCID: PMC7907684.

4. Favril L, Yu R, Hawton K and Fazel S. Risk factors for self-harm in prison: a systematic review and meta-analysis, The Lancet Psychiatry. 2020;7(8) pp682–91.

5. Lanes, E. Identification of Risk Factors for Self-Injurious Behavior in Male Prisoners, J Forensic Sci, May 2009, 54(3) pp692–698 doi: 10.1111/j.1556-4029.2009.01028.x

6. Ryland H, Gould C, McGeorge T, Hawton K and Fazel S. Predicting self-harm in prisoners: Risk factors and a prognostic model in a cohort of 542 prison entrants, European Psychiatry, 2020;28;63(1):e42. doi: 10.1192/j.eurpsy.2020.40.

7. Stephenson T, Leaman J, O’Moore É, Tran A and Plugge E. Time out of cell and time in purposeful activity and adverse mental health outcomes amongst people in prison: a literature review. Int J Prison Health, 2021 6;17(1) pp54–68. doi: 10.1108/IJPH-06-2020-0037. PMID: 33634654.

8. Hewson T, Gutridge K, Bernard Z, Kay K and Robinson L. A systematic review and mixed-methods synthesis of the experiences, perceptions and attitudes of prison staff regarding adult prisoners who self-harm. BJPsych Open, 2022:8(4), E102. doi: 10.1192/bjo.2022.70. PMID: 35659128; PMCID: PMC9230562.

9. Borrill J, Snow L, Medlicott D, Teers R and Paton J. “Learning from ‘near misses’: interviews with women who survived an incident of severe self-harm in prison”, The Howard Journal of Criminal Justice, 2005, 44(1), pp. 57–69.

10. Marzano L, Fazel S, Rivlin A and Hawton K, “Near-lethal self-harm in women prisoners: contributing factors and psychological processes”, Journal of Forensic Psychiatry & Psychology, 2011, 22(6), pp. 863–884.

11. Baggio S, Getaz L, Tran NT, Peigne N and Chacowry Pala K. Association of overcrowding and turnover with self-harm in a Swiss pre-trial prison, International Journal of Environmental Research and Public Health, 2018, 15(4) pE601.

12. Walker T, Shaw J, Gibb J, Turpin C, Reid C, Gutridge K and Abel K (2021) Lessons learnt from the narratives of women who self-harm in prison, Crisis, 42(4) p255–262.

13. Facer-Irwin, E, Blackwood, N, Bird, A and MacManus, D.Trauma, post-traumatic stress disorder and violence in the prison population: Prospective cohort study of sentenced male prisoners in the UK. BJPsych Open. 2023;9(2), E47. doi:10.1192/bjo.2022.639

14. National Institute for Health and Care Excellence. Self-harm: assessment, management and preventing recurrence, NICE guideline 225. 2022 [Available online at https://www.nice.org.uk/guidance/ng225, accessed 11/10/23.]

15. Nock, MK, Holmberg, EB, Photos, VI, & Michel, BD. The Self-Injurious Thoughts and Behaviors Interview: Development, reliability, and validity in an adolescent sample. Psychological Assessment, 2007;19 pp309– 317.

16. HM Inspectorate of Prisons a. Report on an unannounced inspection of HMP Wandsworth by HM Chief Inspector of Prisons (13 and 20-24 September 2021) - further resources: Prisoner survey methodology, results and analyses HMP Wandsworth September 2021. 2021 [Available online at https://www.justiceinspectorates.gov.uk/hmiprisons/inspections/hmp-wandsworth-3/, accessed 15/11/23.]

17. HM Inspectorate of Prisons b. Report on an unannounced inspection of HMP Wandsworth by HM Chief Inspector of Prisons (13 and 20-24 September 2021) - further resources: Population statistics. 2021 [Available online at https://www.justiceinspectorates.gov.uk/hmiprisons/inspections/hmp-wandsworth-3/, accessed 15/11/23.]

18. Kaba F, Lewis A, Glowa-Kollisch S, Hadler H, Lee D, Alper H, Selling D, MacDonald R, Solimo A, Parsons A and Venters H. Solitary Confinement and Risk of Self-Harm Among Jail Inmates, American Journal of Public Health 2014;104(3) pp442–447.

19. Martin MS, Dorken SK, Colman I, McKenzie K and Simpson AIF. The incidence and prediction of self-injury among sentenced prisoners, Canadian Journal of Psychiatry 2014;59(5) pp259-267.

20. Vinokur D and Levine SZ. Non-suicidal self-harm in prison : A national population-based study, Psychiatry Research 2019;272 pp216–221 doi: 10.1016/j.psychres.2018.12.103. Epub 2018 Dec 19. PMID: 30590275.

21. Smith H and Kaminski R. Inmate Self-injurious behaviours: Distinguishing characteristics within a retrospective study, Criminal Justice and behavior 2010;37(1) pp81–96.

22. Leese M, Thomas S and Snow L. An ecological study of factors associated with rates of self-inflicted death in prisons in England and Wales. Int J Law Psychiatry 2006 Sep-Oct;29(5) pp355–60. doi: 10.1016/j.ijlp.2005.10.004. Epub 2006 Jun 14. PMID: 16780949.

