## Supplementary material for "Environmental risk factors for self-harm during imprisonment: a prospective cohort study": S1 Baseline questionnaire

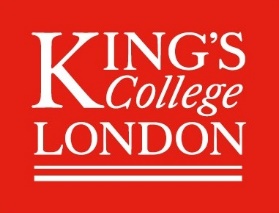

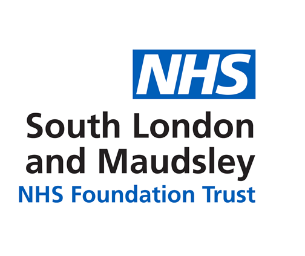

SHAPE study questionnaire

| **Q** | **QUESTION** | | | | | | | | | | | **T** | **S** |
| --- | --- | --- | --- | --- | --- | --- | --- | --- | --- | --- | --- | --- | --- |
| 1 | What is your age? | _______ Years | | | | | | | | | | SD | F |
| 2 | Did you have a job at the point you were arrested? | □ Yes  □ No | | | | | | | | | | SD | F |
| 3 | Have you experienced homelessness in the past (for a month or more)? | □ Yes  □ No | | | | | | | | | | SD | O |
| 4 | Have you been in prison before? | □ Yes  □ No | | | | | | | | | | CR | O |
| 5 | Have you ever been convicted of a violent offence? (This includes offences such as battery, assault, ABH and GBH) | □ Yes  □ No | | | | | | | | | | CR | F |
| 6 | Do you have a court date(s) in the next month? | □ Yes  □ No | | | | | | | | | | CR | N |
| 7 | What is your legal status here in prison? | □ Remand  □ Sentenced  □ Probation license recall  □ Other (e.g. extradition) | | | | | | | | | | CR | F |
| 8 | If you are in prison due to a sentence, or recalled on probation license, what is the length of your sentence or recall period? | _____ Years _____ Months _____ Days | | | | | | | | | | CR | F |
| 9 | Are you in regular contact with any friends or family members outside of the prison? (For example, over the phone or by in-person or video visit.) | □ Yes  □ No | | | | | | | | | | E | N |
| 10 | If you are not in regular contact with friends or family outside prison, what is the main reason for this? | □ My PIN has not yet been approved  □ I have not yet added the relevant numbers to my PIN  □ I am not yet permitted an in-person/video visit  □ Another reason ____________ (please state)  □ I’d prefer not to say | | | | | | | | | | E | N |
| 11 | Do you use a vape in prison? | □ Yes  □ No | | | | | | | | | | E | N |
| 12 | Were you ever in local authority care before the age of 16? | □ Yes  □ No | | | | | | | | | | H | O |
| 13 | At any age, have you ever been the victim of sexual abuse? | □ Yes  □ No | | | | | | | | | | H | F |
| 14 | Have any of your family died by suicide or self-harmed themselves? | □ Yes  □ No | | | | | | | | | | H | O |
| 15 | Are you currently diagnosed with an emotional or psychiatric disorder? | □ Yes  □ No | | | | | | | | | | C | Ob |
| 16 | Are you currently prescribed any medication for emotional, psychological or psychiatric problems? | □ Yes  □ No | | | | | | | | | | C | F  Ob |
| 17 | Have you ever had psychiatric treatment by a medical health professional? | □ Yes  □ No | | | | | | | | | | C | O |
| 18 | Have you ever had psychiatric treatment whilst in prison? | □ Yes  □ No | | | | | | | | | | C | F |
| 19 | Have you ever experienced suicidal thoughts during your life? (Choose the answer which fits best) | □ Yes, in the last month  □ Yes, but not in the last month  □ Yes, but not in the last year  □ No, never | | | | | | | | | | C | F |
| 20 | Have you ever self-harmed outside of prison? | □ Yes  □ No | | | | | | | | | | C | Ob |
| 21 | Have you ever attempted suicide outside prison? | □ Yes  □ No | | | | | | | | | | C | Ob |
| 22 | Have you ever self-harmed inside of prison? | □ Yes  □ No | | | | | | | | | | C | Ob |
| 23 | Have you ever attempted suicide inside prison? | □ Yes  □ No | | | | | | | | | | C | Ob |
| 24 | Do you have current thoughts about wanting to harm yourself? | □ Yes  □ No | | | | | | | | | | C | O |
| 25 | Could you describe the most stressful event that has occurred for you in the past week? | ___________________________________  ___________________________________  _____________________ (please describe) | | | | | | | | | | E | N |
| 26 | How distressing was this event for you? | Not at all 🡨 | | |  | |  | | 🡪 Totally | | | E | N |
|  |  | 1 | 2 | 3 | | 4 | | 5 | | 6 | 7 |  |  |
| 27 | Do you feel that the future is hopeless and that things cannot improve? | □ Yes  □ No | | | | | | | | | | C | O |
| 28 | Do you have any long-term health conditions? If yes, please name or describe it/them | □ Yes  □ No  Name/description of condition(s) ________________________________ | | | | | | | | | | C | F |
| 29 | Were you misusing alcohol (drinking alcohol to excess) before coming into prison? | □ Yes  □ No | | | | | | | | | | C | F |
| 30 | Were you using drugs before coming into prison? | □ Yes  □ No | | | | | | | | | | C | F |
|  | *If yes to Q30, please go to questionnaire 2 (DUDIT). If no, go to questionnaire 3 (CTQ).* | | | | | | | | | | |  |  |
| SHAPE study questionnaire v4.0 24.06.22 IRAS 306528  T = Variable type, S = Variable source, SD = sociodemographic, CR = criminological, E = environmental, H = historical, C = clinical, F = Favril et al., O = OxSHIP, N = New item | | | | | | | | | | | | | |

S1 Supplementary figure 1. 30-item study questionnaire.
