## Supplementary material for "Environmental risk factors for self-harm during imprisonment: a prospective cohort study": S2 Exit questionnaire

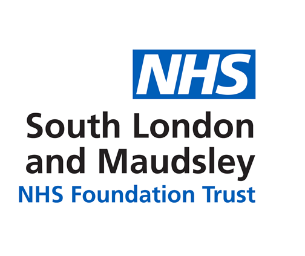

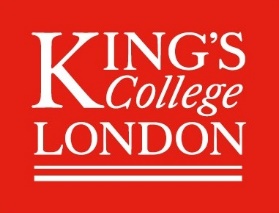

SHAPE study exit questionnaire

We are asking you to answer these extra questions after spending three months in HMP Wandsworth. This is because we think they are important but the answers are not easy to find in your computer records.

The same confidentiality as before (in your original questionnaires) applies to these questions. Your rights regarding your personal information remain the same as before (see Participant Information Sheet for more details).

| **Q** | **QUESTION** | | **T** | **S** |
| --- | --- | --- | --- | --- |
| 1 | Since you started your current spell at HMP Wandsworth, how many different cellmates have you had? | ______ number of cellmates | E | N |
|  | **Thoughts of non-suicidal self-injury** | | | |
| 2 | Since you started your current spell at HMP Wandsworth, have you ever had thoughts of purposely hurting yourself without wanting to die?  *(for example, cutting or burning)* | □ Yes  □ No | O | N |
|  | *We will refer to this as* ***non-suicidal self-injury*** | |  |  |
| 3 | How many separate times in the past three months? | ________________ (number of times) | O | N |
| 4 | How many separate times in the past week? | ________________ (number of times) | O | N |
|  | **Non-suicidal self-injury** | | | |
| 5 | Since you started your current spell at HMP Wandsworth, have you ever actually engaged in NSSI? | □ Yes  □ No | O | N |
| 6 | How many separate times in the past three months? | ________________ (number of times) | O | N |
| 7 | How many separate times in the past week? | ________________ (number of times) | O | N |
| **End of questions. Thank you for your time.** | | | | |

S2 Supplementary figure 2. 5-item exit questionnaire.
