## Supplementary table 1 for "Environmental risk factors for self-harm during imprisonment: a prospective cohort study"

| **Outcome** | **Assessment method** | **Count positive/total (%)^1^** |
| --- | --- | --- |
| Self-harm behaviour | Medical records | 14/82 (17.1) |
|  | Self-report at end of follow-up | 13/78 (16.7) |
| Self-harm ideation | Self-report at end of follow-up | 22/78 (28.2) |
| ^1^ All counts after excluding one participant placed in prison healthcare wing | | |

S3 Supplementary table 1. Outcome event rates.
